# Personalized Hemodynamic Management Using Reinforcement Learning to Prevent Persistent Acute Kidney Injury After Cardiac Surgery

**DOI:** 10.1101/2025.10.23.25338698

**Authors:** Moein Sabounchi, Jacob M Desman, Idan Shenfeld Amit, Wonsuk Oh, Chris Capone, Pushkala Jayaraman, Gagan Kumar, Michelle Campoli, Mahima Vijayaraghavan, Prem Timsina, Paul McCarthy, Anthony Manasia, John Oropello, Robin Varghese, Ksenia Gorbenko, Hernando Gomez-Danies, Patricia Kovatch, Gordon Smith, Avniel Shetreat-Klein, Ashita Tolwani, Mayte Suárez-Fariñas, Kianoush Kashani, Ashish Khanna, Azra Bihorac, Jolion McGreevy, Lisa Stump, John Kellum, David Reich, Pulkit Agrawal, Girish N Nadkarni, Ankit Sakhuja

**Affiliations:** Charles Bronfman Institute for Personalized Medicine, Icahn School of Medicine at Mount Sinai, New York, New York, USA; Windreich Department of Artificial Intelligence and Human Health, Icahn School of Medicine at Mount Sinai, New York, New York, USA; Improbable AI Lab, Massachusetts Institute of Technology, Cambridge, Massachusetts, USA; Department of Pulmonary and Critical Care Medicine, Northeast Georgia Medical Center, Gainesville, Georgia, USA; Division of Hospital Medicine, Department of Medicine, Icahn School of Medicine at Mount Sinai, New York, New York, USA; Clinical Data Science, Mount Sinai Hospital, New York, New York, USA; Section of Cardiovascular Critical Care, Department of Cardiovascular and Thoracic Surgery, West Virginia University, Morgantown, West Virginia, USA; Institute for Critical Care Medicine, Icahn School of Medicine at Mount Sinai, New York, New York, USA; Department of Cardiothoracic Surgery, Icahn School of Medicine at Mount Sinai, New York, New York, USA; Department of Population Health Science and Policy, Icahn School of Medicine at Mount Sinai, New York, New York, USA; Department of Emergency Medicine, University of Pittsburgh School of Medicine, Pittsburgh, Pennsylvania, USA; Scientific Computing, Icahn School of Medicine at Mount Sinai, New York, New York, USA; Department of Epidemiology and Biostatistics, West Virginia University, Morgantown, West Virginia, USA; Department of Rehabilitation and Physical Medicine, Icahn School of Medicine at Mount Sinai, New York, New York, USA; Division of Nephrology, Department of Medicine, University of Alabama at Birmingham, Birmingham, Alabama, USA; Division of Nephrology and Hypertension, Division of Pulmonary and Critical Care Medicine, Department of Medicine, Mayo Clinic, Rochester, Minnesota, USA; Department of Anesthesiology, Division of Critical Care Medicine, Atrium Health Wake Forest Baptist Medical Center, Wake Forest School of Medicine, Winston-Salem, North Carolina, USA; Outcomes Research Consortium, Houston, Texas, USA; Division of Nephrology, Hypertension, and Renal Transplantation, Department of Medicine, College of Medicine, University of Florida, Gainesville, Florida, USA; Intelligent Clinical Care Center, The University of Florida College of Medicine, Gainesville, Florida, USA; Department of Emergency Medicine, Icahn School of Medicine at Mount Sinai, New York, New York, USA; Mount Sinai Health System and Icahn School of Medicine at Mount Sinai, New York, New York, USA; Department of Critical Care Medicine, University of Pittsburgh School of Medicine, Pittsburgh, Pennsylvania, USA; The Hasso Plattner Institute for Digital Health at Mount Sinai, Icahn School of Medicine at Mount Sinai, New York, New York, USA

## Abstract

**Importance:** Acute kidney injury (AKI) affects one-third of patients after cardiac surgery and increases morbidity and mortality. AKI lasting over 48 hours, known as persistent AKI (pAKI), has much worse outcomes. Hemodynamic optimization is cornerstone of AKI management, however, current strategies rely on bundled care interventions that are inconsistently implemented, underscoring the need for personalized hemodynamic optimization.

**Objective:** To develop and validate a reinforcement learning (RL) model to guide individualized dosing of intravenous (IV) fluids, vasopressors, and inotropes for prevention of pAKI after cardiac surgery.

**Design:** Cohort study. Model development and internal validation were performed retrospectively in MIMIC-IV, with external validation in SICdb, a European database (retrospective), and then in Mount Sinai Health System cohort using data from Jan 1–Aug 18, 2025).

**Setting:** Multicenter retrospective cohort study.

**Participants:** Admissions to ICU after cardiac surgery.

**Exposures:** Postoperative hemodynamic management during first 72 hours of ICU stay using IV fluids, vasopressors, and inotropes.

**Main Outcomes and Measures:** Primary outcome was pAKI within 5 days after surgery. The RL model optimized treatment policies through reward-based learning, where higher rewards reflected improved outcome. We assessed model performance relative to clinicians using Fitted Q Evaluation and adjusted weighted pooled logistic regression.

**Results:** There were 6,643 adult ICU admissions following cardiac surgery in MIMIC-IV, 2,254 in SICdb, and 846 in MSHS. Median age was 70 years in MIMIC-IV, 70.0 years in SICdb, and 64 years in MSHS cohort with 72%, 73%, and 70% males respectively. AKI occurred in 41.4%, 19.7%, and 22.5% of admissions, with pAKI in 30.5%, 43.0%, and 33.7% of AKI cases, respectively. RL model achieved higher cumulative rewards than clinicians across all cohorts. Concordance between clinician actions and RL model’s recommendations was associated with lower adjusted odds of pAKI (OR, 0.92 [0.89–0.96] in SICdb; 0.91 [0.86–0.96] in MSHS). RL model favored smaller IV fluid volumes, moderate vasopressor dosing, and greater inotrope use.

**Conclusions and Relevance:** In this study, personalization of early postoperative hemodynamic management using an RL model was associated with decreased risk of pAKI. These findings suggest that AI guided hemodynamic strategies may enhance postoperative care after cardiac surgery.

**Key Points:** *Question:* Can reinforcement learning (RL) personalize early postoperative hemodynamic management to prevent persistent AKI (pAKI) after cardiac surgery?

*Findings:* In 9,743 postoperative cardiac surgery ICU admissions across 3 cohorts (MIMIC-IV, SICdb, and Mount Sinai Health System), the RL model achieved higher cumulative rewards than clinician policies and was associated with lower adjusted odds of developing pAKI when clinician actions aligned with model recommendations. The RL model favored smaller intravenous fluid volumes and earlier, graded adjustments in vasopressor and inotrope dosing compared with standard practice.

*Meaning:* RL guided individualized hemodynamic management after cardiac surgery shows promise in reducing the risk of persistent AKI and should be tested in randomized clinical trials.

## INTRODUCTION

Cardiac surgery is common and involves significant physiological stress and hemodynamic instability in the early postoperative period. Acute kidney injury (AKI),^1^ defined by a rise in serum creatinine occurs in over one-third of patients following cardiac surgery and associated with a four-fold increase in mortality and doubling of health-care costs.^2–4^ Outcomes are substantially worse when AKI persists for over 48 hours ^5^, also known as persistent AKI (pAKI).^6^

The management of AKI is largely supportive with hemodynamic optimization as its cornerstone. This is especially important in the early post-operative period after cardiac surgery when patients have a dynamic hemodynamic profile that requires simultaneous use of intravenous (IV) fluids, vasopressors and inotropes. Although these interventions when used as part of a clinical bundle have shown benefit,^7^ their uptake in routine practice remains very poor.^8,9^

pAKI itself is a heterogeneous syndrome, with distinct phenotypes that exhibit variable responses to therapy.^10^ Even when phenotypes are defined by widely available clinical data rather than specialized biomarkers, differences in disease trajectory and treatment response persist.^11^ These findings highlight the need for personalized strategies to guide AKI management, particularly among high-risk cardiac surgery patients.

Reinforcement learning^12^ (RL), an AI approach that optimizes sequential decision-making, offers a promising framework for this challenge. In RL, the model learns to make decisions by taking actions in an environment and receiving feedback in the form of rewards or penalties.^13^ This iterative learning process enables the agent to develop policies that maximize cumulative rewards over time. In healthcare, where clinical trajectories evolve dynamically, RL is particularly well-suited for guiding time-sensitive interventions such as titration of IV fluids, vasopressors, and inotropes in response to continuously changing physiological states. Prior applications of RL in critical care have shown promise^14,15^, including recent work demonstrating its utility for postoperative glucose control in cardiac surgery patients.^16^ Building on this foundation we introduce an RL model designed to guide early postoperative hemodynamic management in order to prevent pAKI following cardiac surgery.

## METHODS

### Study Databases and Population

We used data from adult (≥18 years) patients who had postoperative cardiac surgery intensive care unit (ICU) admissions in the Medical Information Mart for Intensive Care (MIMIC-IV) database as the development cohort and performed external validation in the retrospective cohort from Salzburg Intensive Care Database (SICdb)^17^ and data from Jan 1, 2025 to Aug 18 2025 from the Mount Sinai Health System (MSHS). Ethical approval was obtained from the Icahn School of Medicine at Mount Sinai Institutional Review Board (study number 20-00338). The overall structure of study is shown in Figure 1. We excluded admissions with end-stage kidney disease and those with missing data for serum creatinine, weight, or IV fluids. As the model was trained on data from the first 72 hours of ICU stay, patients who did not survive beyond this period were excluded from retrospective cohorts to avoid bias from nonrepresentative clinical trajectories. We have shared the details of the datasets and inclusion/exclusion criteria in e-Methods, e-Table 1 and e-Figure 1 in the Supplement.

**Fig. 1.**
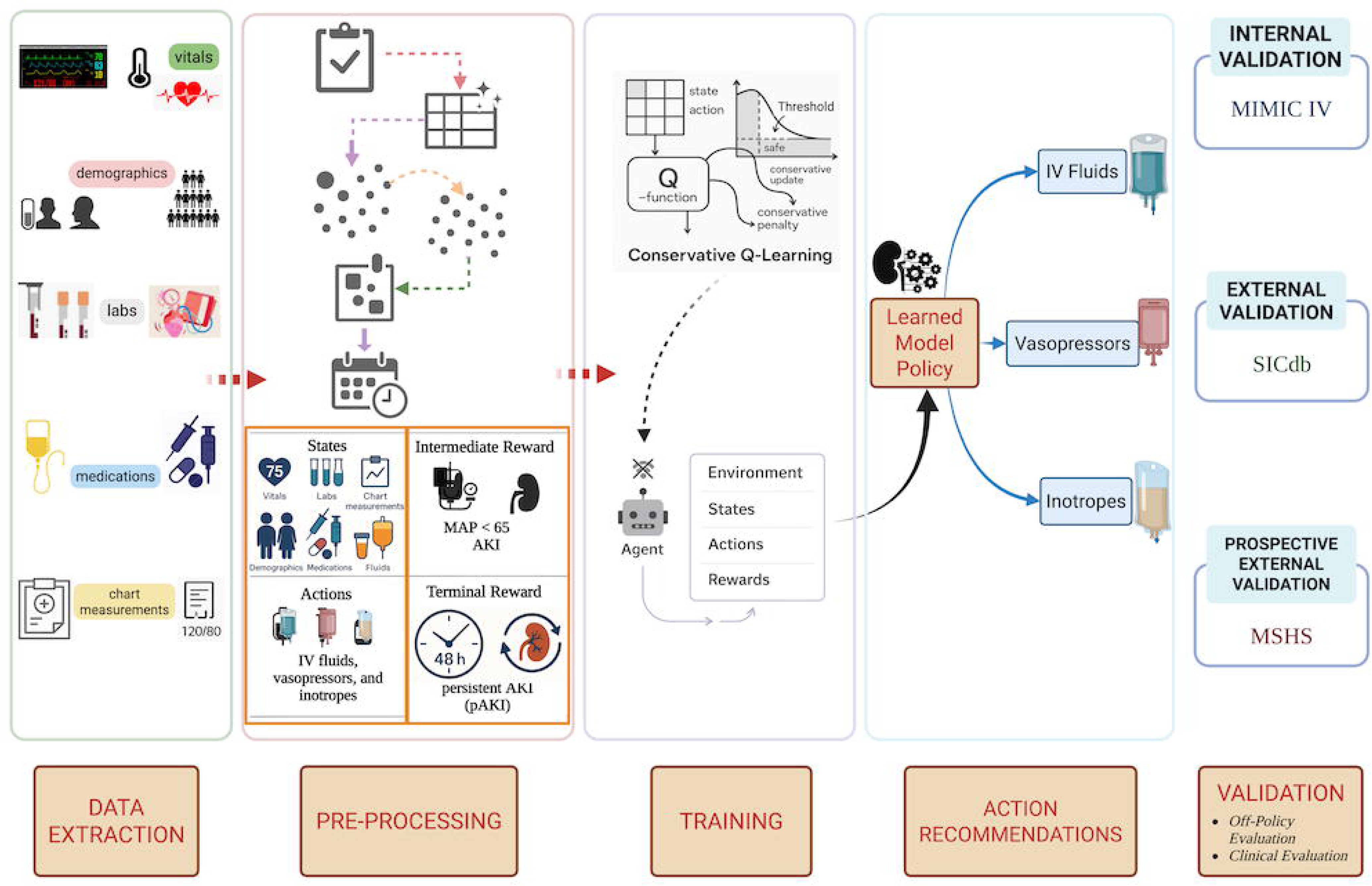
Overview of the Study **Abbreviations:** IV Fluids, Intravenous Fluids; MAP, Mean Arterial Pressure; AKI, Acute Kidney Injury; MIMIC IV, Medical Information Mart for Intensive Care (MIMIC)-IV; SICdb, Salzburg Intensive Care database; MSHS: Mount Sinai Hospital System.

### Feature Extraction and Preprocessing

We extracted features routinely available to clinicians at bedside for development of the RL model. These included demographics (age, sex), anthropometrics (height, weight, BMI), vital signs, laboratory values, fluid balance, medications (vasopressors, inotropes, IV fluids including crystalloids and colloids, and nephrotoxins), and SOFA Score. We converted the vasopressor and inotrope doses into their norepinephrine (NEE) and dobutamine equivalent (DE) doses^18,19^ respectively.

Data were extracted from ICU admission up to the earlier of either 72 hours or ICU discharge. We removed biologically implausible values.^20^ The resulting time-series data were segmented into hourly bins, with features either summed or averaged as clinically appropriate. We excluded features with more than 30% missingness. We imputed missing values using established practices, using a combination of forward filling and k-nearest neighbor imputation.^16^

### Outcome

The primary outcome of our study was pAKI within 5 days after cardiac surgery. We defined pAKI as AKI lasting for 48 hours or longer.^6^ We defined AKI as per the Kidney Disease Improving Global Outcomes (KDIGO) guidelines as an increase in serum creatinine by 0.3mg/dL or more within 48 hours or an increase by at least 1.5 times the baseline serum creatinine within 7 days.^21^ We used the last available pre-operative creatinine within the five days before surgery as the baseline creatinine.^22^ If no pre-operative value was available, we used the first post-operative creatinine instead.^22^

### Model Training

We used conservative Q-learning (CQL)^23^ as the RL algorithm. We specified states, actions and rewards for the RL model on an hourly basis. States included laboratory values, vital signs, medications (IV fluids, vasopressors, inotropes, nephrotoxins), demographics, anthropometrics (height, weight, BMI), SOFA score.^24^ We defined actions as recommended doses of IV fluids, inotropes and vasopressors discretized into clinically meaningful categories (e-Table 2). We calculated rewards for each epoch as a weighted combination of pAKI, AKI and MAP < 65mmHg. We have provided a details of computational modeling, state, actions and reward formulations in e-Methods in the Supplement. We developed the RL model using data from MIMIC-IV which was split into three subsets: 70% training, 15% validation, and 15% internal testing. We then externally validated it in the retrospective SICdb (and prospective MSHS cohorts.

During training we used a discount factor rate of 0.99 which emphasizes long-term rewards and reflects the clinical importance of avoiding adverse outcomes over time. We set the conservativeness coefficient (alpha) to 0.25 which controls the degree to which the learned policy diverges from the observed clinician behavior, encouraging safe exploration without straying too far from established practices. We trained the model using a batch size of 512, an initial learning of 0.001 with a learning rate scheduler that progressively reduced the rate to 1e-8.

### Statistical Analysis

We described continuous variables as median (inter quartile range; IQR) and categorical variables as counts (percentages). We used Kruskal-Wallis test ^25^ for continuous variables and Chi-squared test^26^ categorical variables. We set the significance level at 0.05 without adjusting for multiple comparisons.

We first evaluated the model’s performance using Fitted Q evaluation (FQE) ^27^, an established OPE method to compare total rewards of an RL model to that of clinicians in the internal test set. To assess generalizability, we then used FQE to assess the performance of the model in retrospective SICdb and prospective MSHS cohorts.

Next, we conducted a clinical evaluation comparing model’s recommendations with clinician actions to assess how the model’s learned policy differed from observed clinical practice. We examined how recommendations evolved over time and across mean arterial pressure (MAP) thresholds. To assess patient-level patterns, we randomly sampled 30 admissions from each external cohort, including 10 with pAKI, 10 with AKI only, and 10 without AKI, and generated heatmaps of mean arterial pressure trajectories, clinician actions, and model’s hourly recommendations.

Finally, we used a weighted pooled logistic regression^28,29^ to estimate the odds of development of pAKI when clinician actions were concordant with model’s recommendations. To account for time-varying confounding, we applied inverse probability of treatment weighting (IPTW) using the same baseline and time-dependent covariates as in model training^28,29^. As standard error estimates can be anti-conservative when using IPTW, we also report results using robust sandwich variance estimator^29–31^.

### Interpretability

Interpretability is essential in RL for clinical decision-making to ensure transparency and clinician trust, particularly when recommending high-risk interventions. We used SHAP (SHapley Additive exPlanations) to assess both global and local interpretability of the RL model ^32^. We selected SHAP for its strong theoretical foundation and ability to quantify the average contribution of each input feature to the model’s overall predictions. Shapley analysis stems from cooperative games in game theory and it is used to fairly distribute the payoffs in cooperative game to each contributing player. In RL, SHAP value represent the relative influence of each feature on the model’s recommended actions.^16^ We computed SHAP values across the entire test set to identify the most influential clinical variables and evaluate their consistency with clinical expectations. We also computed hourly, patient-specific SHAP values to visualize how feature importance evolved over time.

### Model Fairness

To evaluate model robustness across demographic subgroups, we assessed model’s performance using off-policy evaluation and concordance analyses stratified by sex and race.

## RESULTS

### Patient Characteristics

We included 6,643 postoperative cardiac surgery ICU admissions from MIMIC-IV database which served as the development cohort. We externally validated on the performance of the RL model on 2,254 admissions from the retrospective SICdb dataset and 846 admissions from the prospective MSHS cohort. The median age was 69.6 (61.6, 77.1) years in MIMIC-IV, 70.0 (60.0, 75.0) years in SICdb and 64.3 (56.1, 72.2) years in MSHS, with 71.9%, 72.9% and 70.6% males respectively. AKI occurred in 2750 (41.4%) of MIMIC-IV, 444 (19.7%) of SICdb and 190 (22.45%) of MSHS admissions, while pAKI developed among 839 (30.5%), 191 (43.0%) and 64 (33.7%) of AKI cases, respectively (Table 1 and e-Table 3).

**Table 1:** Characteristics of ICU admissions after Cardiac Surgery.

| Feature | MIMIC IV<br>Development - | SICdb External -<br>(n=2254) | MSHS External<br>- | p value |
| --- | --- | --- | --- | --- |

|  | **(n=6643)** |  | **(n=846)** |  |
| --- | --- | --- | --- | --- |
| Admission Age, median (IQR), years | 69.6 (61.6, 77.1) | 70.0 (60.0, 75.0) | 64.3 (56.1, 72.2) | < 0.001 |
| Height (CM), median (IQR), cm | 172.9 (165.0, 178.0) | 170.0 (165.0, 175.0) | 170.2 (162.6, 177.8) | < 0.001 |
| Weight, median (IQR), kg | 83.0 (72.0, 95.7) | 80.0 (70.0, 90.0) | 79.2 (67.6, 91.1) | < 0.001 |
| BMI, median (IQR), kg/m<sup>2</sup> | 28.3 (25.0, 32.2) | 26.3 (24.2, 29.4) | 27.4 (24.3, 30.6) | < 0.001 |
| Maximum Sofa, median (IQR) | 6.0 (4.0, 8.0) | 6.0 (6.0, 8.0) | 10.0 (9.0, 12.0) | < 0.001 |
| Gender (Males Number & Percentage) | 4773 (71.85%) | 1642 (72.85%) | 597 (70.57%) | < 0.001 |
| Baseline Creatinine, median (IQR) | 0.90 (0.80, 1.20) | 0.92 (0.80, 1.10) | 0.88 (0.74, 1.09) | < 0.001 |
| IV Fluids Administered – (ml), median (IQR), ml | 22.6 (10.0, 84.2) | 51.0 (6.6, 100.0) | 26.0 (9.0, 116.0) | < 0.001 |
| Inotrope Doses Administered, median (IQR), mcg/kg/min | 0.00 (0.00, 0.01) | 0.00 (0.00, 0.03) | 0.00 (0.00, 0.00) | < 0.001 |
| Vasopressors Doses Administered, median (IQR), mcg/kg/min | 0.00 (0.00, 0.00) | 0.00 (0.00, 0.00) | 0.00 (0.00, 2.50) | < 0.001 |
| Non-zero IV Fluids Administered – (ml), median (IQR), ml | 31.0 (11.0, 105.3) | 55.0 (22.0, 121.7) | 26.0 (16.0, 116.0) | < 0.001 |
| Non-zero Inotrope Doses Administered, | 5.00 (3.52, 7.50) | 1.92 (1.11, 2.82) | 2.50 (2.50, 5.00) | < 0.001 |
| median<br>(IQR),<br>mcg/kg/min |  |  |  |  |
| Non-zero<br>Vasopressors<br>Doses<br>Administered,<br>median<br>(IQR),<br>mcg/kg/min | 0.05 (0.02, 0.09) | 0.06 (0.03, 0.13) | 0.04 (0.02, 0.08) | < 0.001 |

**Abbreviations:** MIMIC IV, Medical Information Mart for Intensive Care (MIMIC)-IV; SICdb, Salzburg Intensive Care database; MSHS: Mount Sinai Hospital System; IQR: Inter Quartile Range; cm, Centimeters; kg, Kilograms; BMI, Body Mass Index; kg/m<sup>2</sup>, Kilograms per Square Meter; SOFA, Sequential Organ Failure Assessment; IV Fluids, Intravenous Fluids; ml, Milliliters; mcg/kg/min, micrograms per kilogram per minute.

### Model Performance

Using FQE^33^ the model achieved higher cumulative rewards than clinician policies across all datasets, indicating consistently superior performance. The RL model achieved a mean overall reward of 53.86 (95% CI: 52.25 – 55.45) vs. 17.49 (95% CI: 16.28 – 18.68) for clinicians in MIMIC-IV internal test set, 77.36 (95% CI: 76.39-7830) vs. 21.33 (95% CI: 20.27 - 22.38) in SICdb and 73.93 (95% CI: 72.39 – 75.42) vs. 29.47 (28.26 - 30.66) in MSHS (Figure 2).

**Fig. 2.**
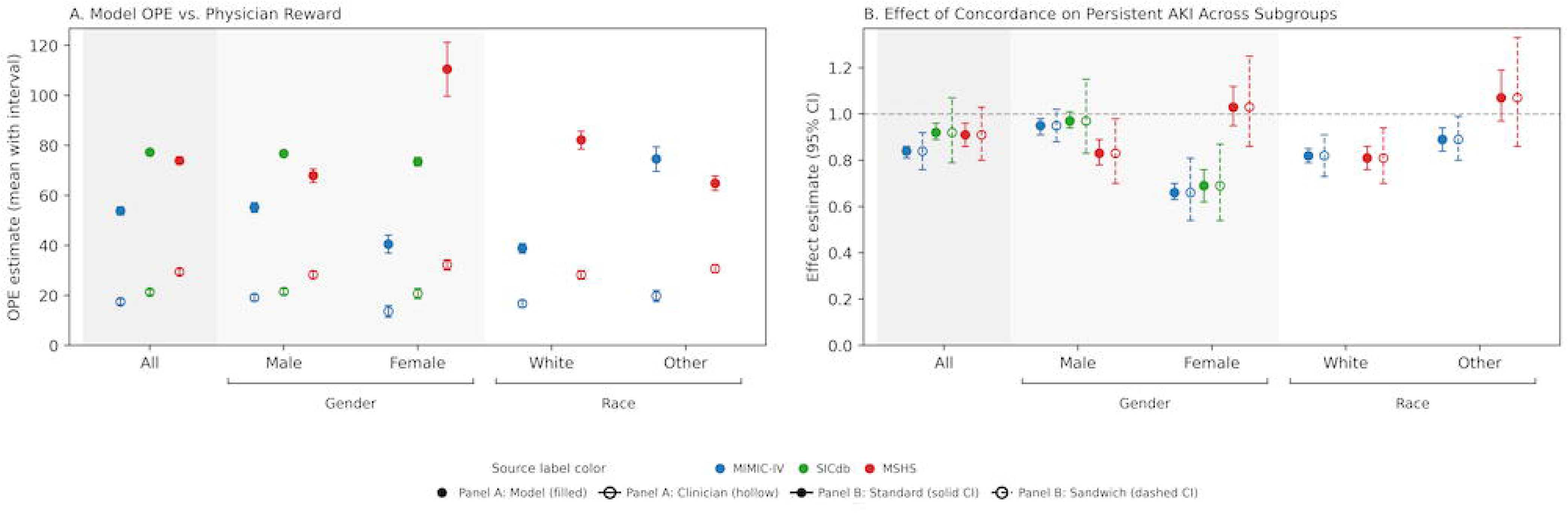
FQE results (a) and Effect of Concordance between Recommendations of the Reinforcement Learning Model and Clinician Actions on Adjusted Odds for Development of Persistent AKI (b) **Abbreviations:** OPE, Off Policy Evaluation; AKI, Acute Kidney Injury; CI: Confidence Interval; MIMIC IV, Medical Information Mart for Intensive Care (MIMIC)-IV; SICdb, Salzburg Intensive Care database; MSHS: Mount Sinai Hospital System.

### Clinical Evaluation of RL Model

Across cohorts, the RL model favored smaller IV fluid volumes, moderate vasopressor doses, and higher inotrope use than clinicians (e-Figure 2). In the SICdb cohort, 0–50 mL IV fluids were recommended by the model in 67.7% of cases vs 30.7% by clinicians (p < .001), and vasopressor dose 0.05–0.1 µg/kg/min NEE in 10.1% vs 7.8% (p < .001) whereas 0.1-0.2 µg/kg/min NEE in 7.9% vs 6.4% (p<0.001). Similar trends were observed in MSHS (IV fluids: 64.9% vs 52.6%, p<0.001; vasopressors: 0.05–0.1 µg/kg/min NEE in 12.3% vs 6.8%, p < .001; 0.1-0.2 µg/kg/min NEE in 6.3% vs 3.5%, p<0.001). The RL model also recommended higher inotrope doses more frequently than clinicians, particularly 5–7.5 µg/kg/min DE (1.4%% vs 0.4% in SiCdb, p < 0.001; 9.7% vs 1.8% in MSHS, p<0.001).

We then evaluated how the model’s dosing policy evolved over time in comparison with clinician actions (e-Figure 2-a). Both clinicians and the model demonstrated temporal shifts in dosing patterns during the first 72 hours after ICU admission. Early in the ICU course, clinicians administered larger volumes of IV fluids with relatively lower doses of vasopressors and inotropes, whereas the model recommended smaller IV fluid volumes but relatively higher doses of vasopressors and inotropes during the same period. In later hours, dosing activity for both clinicians and the model decreased, reflecting a general reduction in therapeutic intensity as patients stabilized.

At MAP < 65 mm Hg, the RL model continued to recommend lower IV fluid volumes (e-Figure 2-b). In the SICdb cohort, the model most frequently recommended 0–50 mL of IV fluids (55.9% vs 28.6% for clinicians, P < .001) and rarely suggested volumes > 500 mL (1.35% vs 10.3%, P < .001). It recommended vasopressor doses 0.05–0.1 µg/kg/min NEE in 16.1% vs 14.1% of cases (P < .001), vasopressor doses 0.1-0.2 µg/kg/min NEE in 15.7% vs 12.7% of cases (P < 0.001) and inotropes of 5–7.5 µg/kg/min DE in 2.8% vs 0.7% (P < .001) cases. In the MSHS cohort, the RL model showed a similar pattern, favoring smaller IV fluid volumes (0–50 mL in 60.9% vs 56.0%, P < .001) and fewer prescriptions > 500 mL (1.7% vs 4.6%, p < 0.001). Vasopressor use followed the same trend, with 0.05 -0.1 µg/kg/min NEE dose in 14.8% vs 8.9% of cases (P < 0.001) and 0.1-0.2 µg/kg/min NEE dose in 7.3% vs 4.2% of cases (P < 0.001). It also more often recommended inotrope doses of 5–7.5 µg/kg/min DE (9.1% vs 0.9% cases, p < .001). Among patients who later developed pAKI, the same overall pattern was observed.

To examine patient-level patterns, we randomly sampled 30 admissions from each external cohort (10 with pAKI, 10 with AKI only, and 10 without AKI) and generated hour-by-hour heatmaps of mean arterial pressure (MAP), clinician actions, and model’s recommendations (Figure 3). Visual comparison of these heatmaps revealed that the RL model generally provided earlier and more graded adjustments in vasoactive support in response to declining MAP. Clinician actions, by contrast, often involved larger, intermittent fluid administrations and delayed or smaller vasoactive adjustments.

**Fig. 3.**
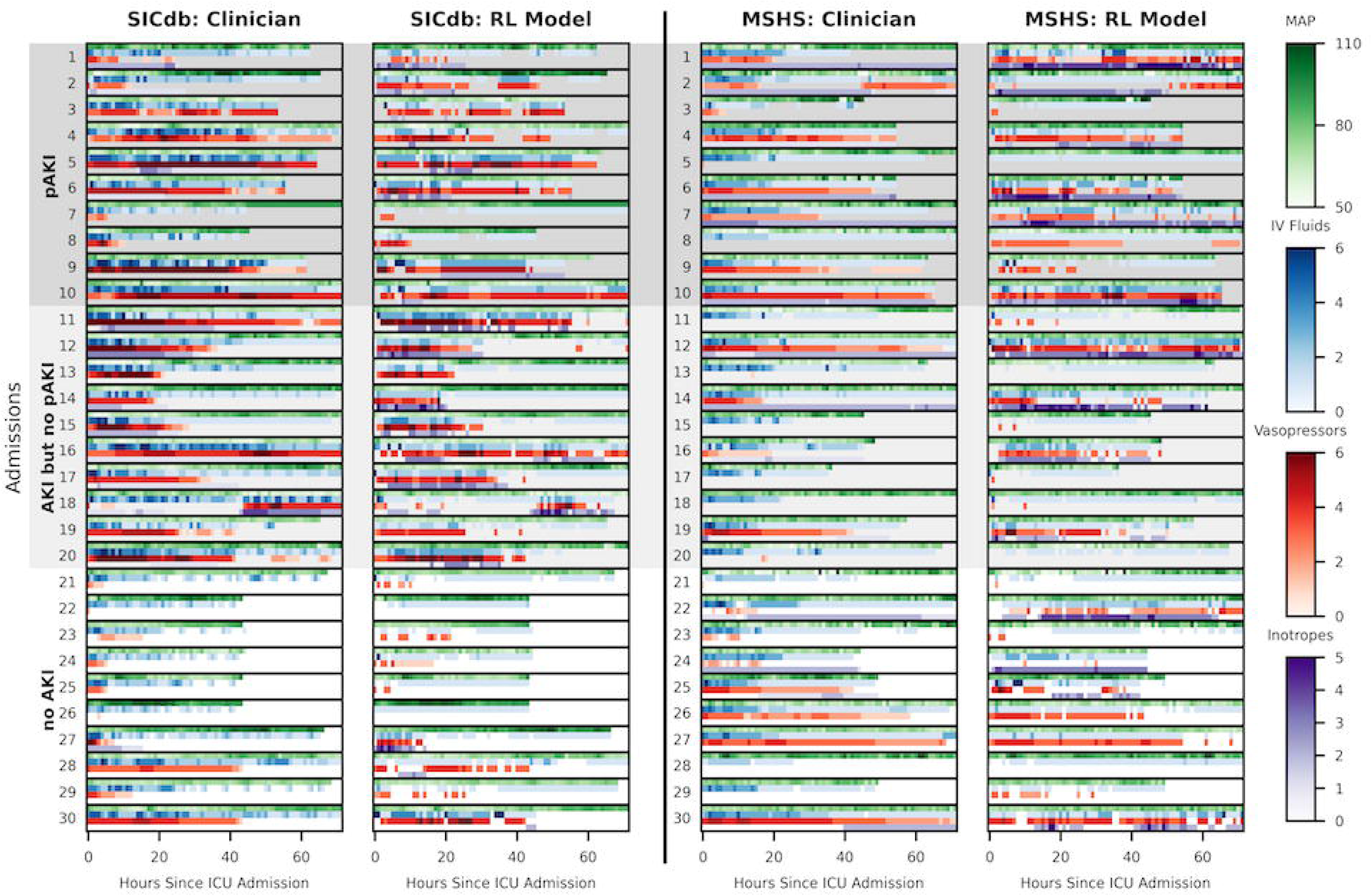
Comparison of Clinician Actions with Recommendations of the Reinforcement Learning Model **Abbreviations:** IV Fluids, Intravenous Fluids; SICdb, Salzburg Intensive Care database; MSHS: Mount Sinai Hospital System; AKI, Acute Kidney Injury; pAKI, persistent Acute Kidney Injury; MAP, Mean Arterial Pressure.

Using weighted pooled multivariable logistic regression with IPTW, concordance between clinician actions and model recommendations was associated with lower odds of pAKI across all cohorts (Figure2) Odds ratios (95% CI) were 0.84 (95% CI: 0.81–0.86) in MIMIC-IV, 0.92 (95% CI: 0.89–0.96) in SICdb, and 0.91 (95% CI: 0.86–0.96) in MSHS using standard variance estimation, and 0.84 (95% CI: 0.76–0.92), 0.92 (95% CI: 0.79–1.07), 0.91 (95% CI: 0.80–1.03) using robust sandwich variance estimator respectively.

### Model Interpretability

To understand the factors driving model’s recommendations, we examined feature importance using SHAP analysis. Global SHAP analysis showed that prior vasoactive actions, SOFA score, creatinine, and fluid balance were the main drivers of model’s dosing recommendations (Figure 4). In SICdb, prior vasopressor actions had the strongest influence, whereas in MSHS, inotrope actions dominated, reflecting cohort-specific treatment patterns. This indicates that the RL model adapts its weighting of features to local practice contexts while preserving consistent physiologic logic across datasets. Local SHAP analyses (e-Figures 3a–j) further demonstrated temporal variation in feature importance within patients, with early decisions influenced by prior vasoactive use and illness severity and later actions shaped by fluid balance and kidney indices.

**Fig. 4.**
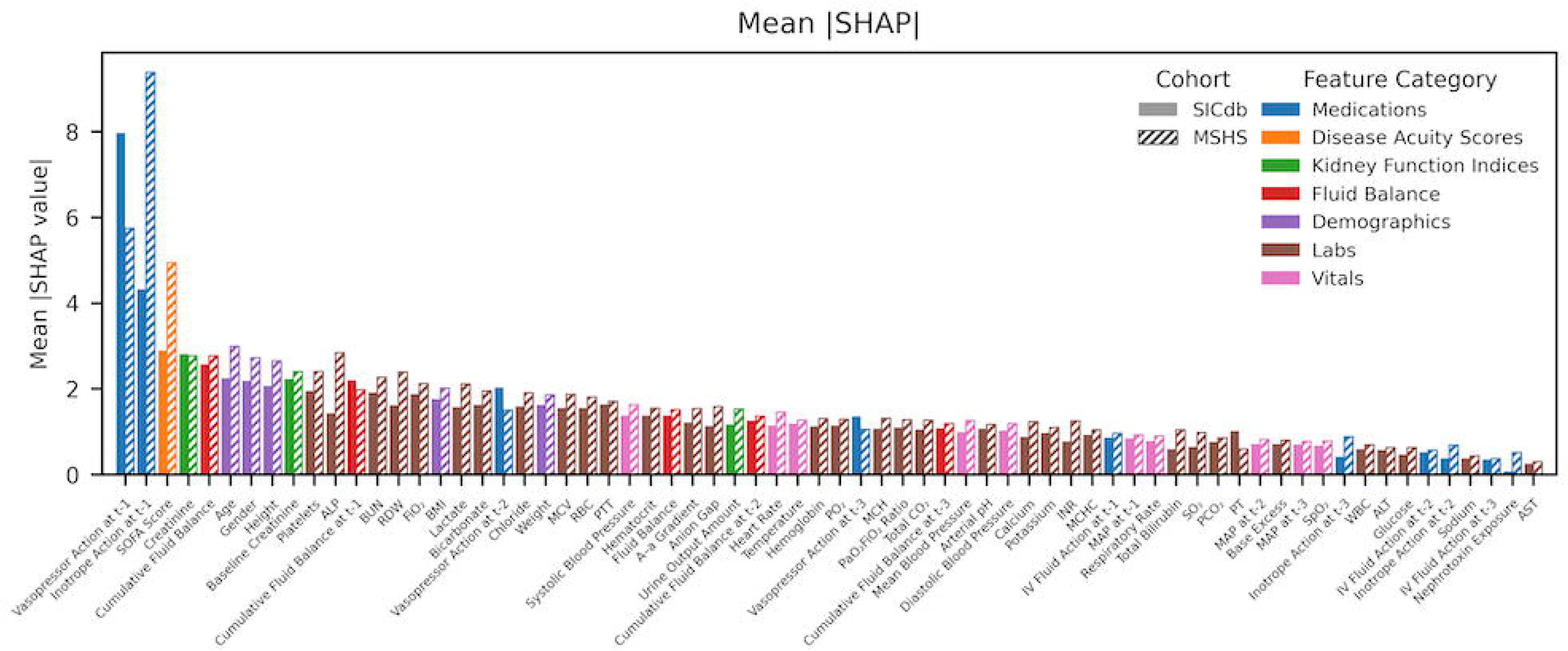
Feature Importance for the Reinforcement Learning Model **Abbreviations:** SHAP, SHapley Additive exPlanations; SICdb, Salzburg Intensive Care database; MSHS: Mount Sinai Hospital System; SOFA, Sequential Organ Failure Assessment; ALP, Alkaline Phosphatase; BUN, Blood Urea Nitrogen; RDW, Red Cell Distribution Width; FiO₂, Fraction of Inspired Oxygen; BMI, Body Mass Index; MCV, Mean Corpuscular Volume; RBC, Red Blood Cell; PTT, Partial Thromboplastin Time; A-a Gradient, Alveolar-arterial oxygen gradient; PO₂, Partial pressure of Oxygen; MCH, Mean corpuscular hemoglobin; PaO₂/FiO₂, Arterial Oxygen Partial Pressure to Fraction of Inspired Oxygen Ratio; CO₂, Carbon Dioxide; Arterial pH, Potential of Hydrogen; INR, International Normalized Ratio; MCHC, Mean Corpuscular Hemoglobin Concentration; IV Fluids, Intravenous Fluids; MAP, Mean Arterial Pressure; SO₂, Oxygen Saturation; PCO₂, Partial Pressure of Carbon Dioxide; PT, Prothrombin Time; SpO₂, Peripheral Capillary Oxygen Saturation; WBC, White Blood Cell; ALT, Alanine Aminotransferase; ; AST, Aspartate Aminotransferase; t-1, previous hour value; t-2, Value in two hours ago; t-3: Value in three hours ago.

### Model Fairness

We evaluated model’s robustness across demographic subgroups using FQE and adjusted concordance analyses stratified by sex and race (Figure 2;). FQE showed that the RL model achieved higher cumulative rewards than clinicians across all subgroups, indicating superior policy performance regardless of sex or race. In multivariable concordance analyses, the adjusted odds ratios for concordance between clinician actions and model recommendations were associated with a lower odds for development of pAKI. No subgroup showed evidence of harm with concordance, demonstrating equitable model performance across demographic groups.

## DISCUSSION

In this multicohort study, we developed and validated an RL model for personalized hemodynamic management in the early post-operative period after cardiac surgery. Across two heterogeneous retrospective cohorts (MIMIC-IV, North America; SICdb, Europe) and a prospective cohort from the Mount Sinai Health System (North America), the RL model consistently achieved higher cumulative rewards than clinician policies and was associated with lower adjusted odds of pAKI when clinician actions aligned with its recommendations.

Cardiac surgery associated AKI remains a leading cause of postoperative morbidity, mortality, and long-term kidney impairment.^2–4^ Outcomes are much worse for patients that develop pAKI.^5,34^ Management of hemodynamics with IV fluids, vasopressors and inotropes is the cornerstone of AKI treatment. However, there is a large variation in their utilization among cardiac surgery patients.^35^ This reflects the underlying heterogeneity of patients, heterogeneity of AKI^10,36^ and differing treatment protocols across institutions. While vasopressors and inotropes are frequently required to maintain perfusion in hemodynamically unstable patients, their use has been associated with higher rates of AKI ^35^, an association that likely reflects underlying disease severity and compromised circulatory status rather than direct nephrotoxicity. Our RL model directly addresses this challenge by personalizing the dosing of IV fluids, vasopressors and inotropes for in the early post-operative phase in these critically ill patients. Moreover, rather than reactively responding to hypotensive events as is common in current standard of care, our RL model recommends proactive, graded adjustments in IV fluids and vasoactive medications aimed at maintaining perfusion and preventing progression to pAKI. This represents a paradigm shift from current standard of care reactive actions to continuous, physiology-informed proactive management of hemodynamics.

Our results align with PrevAKI trial, which demonstrated that patients that received the KDIGO bundle had reduced AKI frequency and severity among cardiac surgery patients.^37^ Importantly, the intervention arm in that trial received more inotropic support compared to usual care. Our model independently converged on a similar pattern, recommending earlier and higher inotrope dosing, particularly when MAPs were declining. This convergence of data-driven strategy and trial-based results strengthen the plausibility of our findings. Our results also align with emerging data that suggests that restrictive fluid strategies after AKI onset are associated higher rates of kidneyrecovery.^38,39^ The fact that our RL model prioritized smaller IV fluid volumes supports this evolving paradigm and suggests that earlier hemodynamic optimization may not equate to aggressive IV fluids administration.

Our study has several strengths. We developed an RL model using a large public ICU database (MIMIC-IV) and externally validated it in two independent cohorts, one retrospective (SICdb) and one prospective (MSHS), supporting generalizability. We used both Fitted Q Evaluation and adjusted concordance analyses to assess policy performance and clinical alignment. Model interpretability analyses identified physiologic variables driving model’s recommendations, reinforcing their clinical plausibility. Subgroup analyses showed generally similar effects across sex and race, with no subgroup demonstrating evidence of harm or systematic bias, addressing a central concern for AI in clinical care.

Our study has some limitations. First, this was an observational study, and only a randomized controlled trial can confirm causal benefit from RL guided therapy. Second, due to lack of consistent availability, we did not include Invasive hemodynamic data such as pulmonary artery catheter measurements. Incorporating these data in future work may further improve model’s performance, however, may decrease generalizability. Finally, we developed this RL model to reduce the development of pAKI within five days after cardiac surgery. While this is a clinically meaningful outcome, future studies should examine longer-term outcomes such as Major Adverse Kidney Events by 30 days.

In summary, we provide a data-driven framework for personalized hemodynamic management after cardiac surgery. By integrating routinely available clinical data, RL can identify personalized strategies that favor IV fluid, vasopressor and inotrope support to prevent the progression to pAKI. Its consistent performance across diverse cohorts highlights the potential for RL to enhance clinician judgment in complex critical care decisions. Future randomized studies are warranted to determine whether implementation of RL guided therapy can improve kidney and patient-centered outcomes.

## Data Availability

The datasets utilized in development and retrospective validation of this RL model can be accessed after completion of necessary training and execution of data use agreement at https://physionet.org/content/mimiciv/2.2/ And: https://www.sicdb.com/
Data from Mount Sinai Health System used for prospective validation is not publicly available.

## Funding

This study was supported by National Institutes of Health (NIH) grant K08DK131286. This work was supported in part through the computational and data resources and staff expertise provided by Scientific Computing and Data at the Icahn School of Medicine at Mount Sinai and supported by the Clinical and Translational Science Awards (CTSA) grant UL1TR004419 from the National Center for Advancing Translational Sciences. The content is solely the responsibility of the authors and does not necessarily represent the official views of the National Institutes of Health.

## COI Statement

This RL model is the subject of a patent cooperation treaty (PCT) application (International Application No. PCT/US2025/047727) in which MS, JMD, GNN, and AS are named inventors. GNN is a founder of Renalytix, Pensieve, Verici and provides consultancy services to AstraZeneca, Reata, Renalytix, Siemens Healthineer and Variant Bio, serves a scientific advisory board member for Renalytix and Pensieve. He also has equity in Renalytix, Pensieve and Verici. AS is a consultant for Roche Diagnostics Corporation. JAK reports receiving consulting fees from Astute Medical/bioMerieux, Astellas, Alexion, Chugai Pharma, Novartis, Mitsubishi Tenabe and GE Healthcare and is a Full-time employee of Spectral Medical. All remaining authors have declared no conflicts of interest.

## Code availability

Code for this study may be shared upon reasonable requests to the corresponding author.

## Author Contributions

Study concept and design: MS, JMD, AS; Acquisition of data: MS, JMD, WO, CC, AS; Analysis and interpretation of data: MS, JMD, ISA, WO, PJ, AS; Drafting of the manuscript: MS, JMD, WO, PJ, AS. Critical revision of the manuscript for important intellectual content: all authors. Statistical analysis: MS, WO; Obtained funding: AS, GNN. Administrative, technical, or material support: AS, GNN; Study supervision: AS

## Notes

### Funding Statement

This study was supported by National Institutes of Health (NIH) grant K08DK131286. The content is solely the responsibility of the authors and does not necessarily represent the official views of the National Institutes of Health.

### Author Declarations

Ethical approval was obtained from the Icahn School of Medicine at Mount Sinai Institutional Review Board (study number 20-00338).

