## Supplement for "Personalized Hemodynamic Management Using Reinforcement Learning to Prevent Persistent Acute Kidney Injury After Cardiac Surgery"

**Supplements**:

**TRIPOD+AI Checklist:**

**eMethods**

**eTable 1-a.** The MIMIC IV (Development) Population cohort ICD Codes

**eTable 1-b.** The MSHS (External Validation) Population Cohort CPT codes

**e-Table 2.** The actions discretization protocol

**e-Table 3:** Patient admissions’ features characteristics: MIMIC IV, SICdb, and MSHS

**e**-**Figure 1:** CONSORT Diagram for inclusion and exclusion criteria of the study

**e**-**Figure 2:** Reinforcement Learning vs. Clinician Actions Comparison

**e**-**Figure 3:** Individual Patients’ SHAP values

**e-Methods:**

**Study Databases and Population:**

MIMIC-IV ^111^ is a publicly available clinical data developed by MIT laboratory for computational Physiology. It contains data from ICUs at Beth Israel Deaconess Medical Center from years 2008-2019. SICdb ^2^ is a European database of 27,000 ICU admissions to the University Hospital Salzburg over 2013-2021 MSHS is a seven hospital health system in New York City. The prospective evaluation of MSHS was performed using data from Jan 1, 2025, to Aug 18^th^ 2025.

In MIMIC-IV, we identified cardiac surgery patients using appropriate ICD-10-PCS codes (e-Table 1a) for coronary artery bypass grafting and/or valve surgery. In SICdb we identified cardiac surgery patients based on the use of "heartsurgerybeginoffset" variable provided in the database ^2^. In MSHS cohort, we included cardiac surgery patients from January to August of 2025 in the Mount Sinai Heath System identified using CPT codes (e-Table 1b).

**Computation Modeling**

We used conservate Q-learning (CQL) ^3^ as the RL algorithm to develop this RL model. CQL is designed to mitigate overestimation of action values in offline settings by penalizing actions not well supported by the training data, thereby reducing the likelihood of unsafe or clinically implausible recommendations ^3^. To further enhance the model’s ability to reason about uncertainty and variability in clinical outcomes, we integrated CQL with a distributional reinforcement learning approach, specifically the Implicit Quantile Network (IQN) ^4^. Unlike standard RL methods that predict only the expected future reward, distributional RL models the entire distribution of possible future outcomes. This richer representation is crucial in clinical decision-making, as it enables the model to account for rare but significant risks, such as acute kidney injury or hypotension. IQN achieves this by learning a quantile-based approximation of the return distribution. Instead of relying on a fixed number of bins, IQN samples quantiles from a continuous distribution, allowing it to flexibly capture a wide range of potential outcomes. This structure not only allows the model to understand the full spectrum of possible future states but also enables fine-tuning of risk preferences via greedy quantile selection. In our implementation, we used 64 sampled quantiles, with 32 designated as greedy quantiles to guide action selection. The combination of CQL and IQN enables this RL model to make more informed and risk-sensitive decisions, especially for interventions with narrow therapeutic indices. Our CQL implementation used an encoder network with four hidden layers of 512 neurons each and ReLU activation function.

RL frames decision-making problems as Markov Decision Processes (MDPs). An MDP is typically defined by a sequence of tuples ($s_{t},a,r,s_{t+1})$ at each time step t, where $s_{t}$ is the observed feature vector representing the state at time t, a is the action taken, and r is the reward received for taking action aa in state $s_{t}$. The resulting state$s_{t+1}$ reflects the system's condition at the next time-step following the action.

**State Space**

The states included features derived from laboratory values, vital signs, medications (vasopressors, inotropes, and nephrotoxins), demographics, anthropometrics (height, weight, BMI), SOFA score and IV fluids. We also included values of mean arterial pressure (MAP) and amount of IV fluids, vasopressors, and inotropes received in the 3-hours prior as part of the state space at time-step t. In total we had sixty-eight states, segmented into one-hour time-windows.

**Action Space**

The actions for the RL model included recommending doses of IV fluids, inotropes and vasopressors discretized into clinically meaningful categories (Supplemental Table 2). The bins were developed based on content expertise of nephrologists and critical care physicians (GK, GNN, AS). We had seven discrete actions for IV fluids and vasopressors each, and six actions for inotropes. This resulted in a total of 294 discrete actions.

**Reward Function**

We designed a clinically informed reward function to guide the agent toward preventing pAKI while maintaining hemodynamic stability. The reward structure is as follows:

a. Terminal Reward:

- If a patient during an admission session develops pAKI in the first 5 days of ICU stay, a terminal penalty of -5 is assigned at the final time-step.
- Conversely, if the patient does not develop pAKI during this time-period, a terminal reward of +5 is given at the time time-step.

b. Intermediate Rewards:

- At any time-step $t$ the model receives a penalty of -2.5 if the patient’s mean arterial pressure falls below 65mmHg at the next step $t+1$.
- At any time-step $t$ if the patient does not meet criteria for AKI, the model receives a reward of +1.

**Concordance analysis methodology:**

We implemented weighted pooled logistic regression to evaluate the effect of concordance, defined as agreement between the RL model’s recommendation and the clinician’s action, on the risk of persistent AKI.

The pooled logistic regression model ^5,6^ was specified as:

$$\mathrm{logit} \left( p\left( Y_{i}=1 \right) \right)=\beta_{0}+\beta_{1}V_{i,0}+\beta_{2}V_{i,t}+\beta_{3}A_{i,t}+f\left( t \right)$$

where $i$ indexes patient, $t$ indexes time (in hour), $Y_{i}$ denotes the binary outcome for persistent AKI, $V_{i,0}$ denotes baseline covariates, $V_{i,t}$ denotes time-dependent covariates, $A_{i,t}$ denotes the concordance indicator between the RL model’s recommendation and the clinician’s action, and $f\left( t \right)$ denotes a natural spline function of time with 1 degree of freedom to flexibly model baseline time trends. Baseline covariates included time independent features such as age and gender, while time-dependent covariates included systolic blood pressure (SBP), diastolic blood pressure (DBP), which used in RL model.

Because both time-dependent covariates and concordance vary hourly, we applied inverse probability of treatment weighting (IPTW) ^5^ to adjust for time-varying confounding. The stabilized IPTW for patient $i$, at time $t$ was defined as:

$$\omega_{i,t}=\prod_{k=0}^{t} \frac{p\left( A_{i,k}=1 | A_{i,k-1},V_{i,0} \right)}{p\left( A_{i,k}=1 | A_{i,k-1},V_{i,0},V_{i,k} \right)}$$

where the numerator model included only baseline covariates and the denominator model included both baseline and time-dependent covariates, consistent with the Vasopressin Initiation RL model. To prevent instability from extreme weights, IPTWs were truncated to the range [0.01, 10].

We estimated odds ratios for concordance effects using both standard model-based (standard) standard errors (SEs) and robust (sandwich)^7,8^ SEs to account for repeated measures. Subgroup analyses were performed by sex (male vs female), and race (White vs other).

**eTable 1-a. The MIMIC IV (Development) Population cohort ICD Codes**

| **Surgery Type** | **Type** | **ICD-10 PCS Code** |
| --- | --- | --- |
| **CABG (Coronary Artery Bypass Grafting)** | Aortocoronary Bypass | 361X |
|  | Coronary Artery (One, two, three, four or more arteries) | 02100X |
|  |  | 02110X |
|  |  | 02120X |
|  |  | 02130X |
| **Valve Repair or Replacement** | Open and other replacement of unspecified heart valve | 352X |
|  | Aortic Valve Replacement, | 02RF0X |
|  | Tricuspid Valve Replacement | 02RG0X |
|  | Pulmonary Valve Replacement | 02RH0X |
|  | Tricuspid Valve Replacement | 02RJ0X |
|  | Aortic Valve Repair | 02QF0X |
|  | Mitral Valve Repair | 02QG0X |
|  | Pulmonary Valve Repair | 02QH0X |
|  | Tricuspid Valve Repair | 02QJ0X |
|  | Open aortic valvuloplasty (Open heart valvuloplasty of aortic valve without replacement) | 3511 |
|  | Open mitral valvuloplasty (Open heart valvuloplasty of mitral valve without replacement) | 3512 |
|  | Open pulmonic valvuloplasty (Open heart valvuloplasty of pulmonary valve without replacement) | 3513 |
|  | Open tricuspid valvuloplasty (Open heart valvuloplasty of tricuspid valve without replacement) | 3514 |

**eTable 1-b. The MSHS (External Validation) Population Cohort CPT codes**

| **Surgery Type** | **Surgery Specification** | **CPT Code** |
| --- | --- | --- |
| **CABG (Coronary Artery Bypass Grafting)** | Coronary Artery Bypass, Vein Only; Single Coronary Venous Graft | 33510 |
|  | Coronary Artery Bypass, Vein Only; 4 Coronary Venous Grafts | 33513 |
|  | Coronary Artery Bypass, Using Venous Graft(S) and Arterial Graft(S); Single Vein Graft (List Separately in Addition to Code for Primary Procedure) | 33517 |
|  | Coronary Artery Bypass, Using Venous Graft(S) and Arterial Graft(S); 2 Venous Grafts (List Separately in Addition to Code for Primary Procedure) | 33518 |
|  | Coronary Artery Bypass, Using Arterial Graft(S); Single Arterial Graft | 33533 |
|  | Coronary Artery Bypass, Using Arterial Graft(S); 2 Coronary Arterial Grafts | 33534 |
|  | Coronary Artery Bypass, Using Arterial Graft(S); 3 Coronary Arterial Grafts | 33535 |
| **Valve Repair or Replacement** | Transcatheter Aortic Valve Replacement (Tavr/Tavi) with Prosthetic Valve; Percutaneous Femoral Artery Approach | 33361 |
|  | Valvuloplasty, Aortic Valve, Open, with Cardiopulmonary Bypass; Complex (Eg, Leaflet Extension, Leaflet Resection, Leaflet Reconstruction, or Annuloplasty) | 33391 |
|  | Replacement, Aortic Valve, Open, with Cardiopulmonary Bypass; with Prosthetic Valve Other Than Homograft or Stentless Valve | 33405 |
|  | Replacement, Aortic Valve; By Translocation of Autologous Pulmonary Valve with Allograft Replacement of Pulmonary Valve (Ross Procedure) | 33413 |
|  | Valvuloplasty, Mitral Valve, with Cardiopulmonary Bypass; with Prosthetic Ring | 33426 |
|  | Valvuloplasty, Mitral Valve, with Cardiopulmonary Bypass; Radical Reconstruction, with or without Ring | 33427 |
|  | Replacement, Mitral Valve, with Cardiopulmonary Bypass | 33430 |
|  | Valvuloplasty, Tricuspid Valve; with Ring Insertion | 33464 |
|  | Replacement, Tricuspid Valve, with Cardiopulmonary Bypass | 33465 |
|  | Replacement, Pulmonary Valve | 33475 |
|  | Ascending Aorta Graft, with Cardiopulmonary Bypass, with Aortic Root Replacement Using Valved Conduit and Coronary Reconstruction (Eg, Bentall) | 33863 |
| **Both/Either** | Reoperation, Coronary Artery Bypass Procedure or Valve Procedure, More Than 1 Month After Original Operation (List Separately in Addition to Code for Primary Procedure) | 33530 |
| **Other Cardiac Surgeries** | Myocardial Resection (Eg, Ventricular Aneurysmectomy) | 33542 |
|  | Ascending Aorta Graft, with Cardiopulmonary Bypass, Includes Valve Suspension, When Performed; for Aortic Dissection | 33858 |
|  | Ascending Aorta Graft, with Cardiopulmonary Bypass with Valve Suspension, with Coronary Reconstruction and Valve-Sparing Aortic Root Remodeling (Eg, David Procedure, Yacoub Procedure) | 33864 |

**Abbreviations:** CPT, Current Procedural Terminology; ICD, International Classification of Diseases.

**e-Table 2.** The actions discretization protocol

| **Discrete Value** | **IV Fluids**  **(ml)** | **Vasopressors**  **(mcg/kg/min)** | **Inotropes**  **(mcg/kg/min)** |
| --- | --- | --- | --- |
| **0** | 0 | 0 | 0 |
| **1** | (0-50] | (0-0.02] | (0-2.5] |
| **2** | (50-100] | (0.02-0.05] | (2.5-5] |
| **3** | (100-250] | (0.05-0.1] | (5-7.5] |
| **4** | (250-500] | (0.1-0.2] | (7.5-10] |
| **5** | (500-1000] | (0.2-0.3] | >10 |
| **6** | > 1000 | > 0.3 | NA |

**Abbreviations**: IV Fluids, Intravenous Fluids; ml, Milliliters; mcg/kg/min, micrograms per kilogram per minute.

**e-Table 3: Patient admissions’ features characteristics: MIMIC IV, SICdb, and MSHS**

| **Feature** | **MIMIC IV Development -**  **(n=6643)** | **SICdb External -**  **(n=2254)** | **MSHS External -**  **(n=846)** | **p value** |
| --- | --- | --- | --- | --- |
| Age, median (IQR), years | 69.6 (61.6, 77.1) | 70.0 (60.0, 75.0) | 64.3 (56.1, 72.2) | < 0.001 |
| Height, median (IQR), cm | 172.9 (165.0, 178.0) | 170.0 (165.0, 175.0) | 170.2 (162.6, 177.8) | < 0.001 |
| Weight, median (IQR), kg | 83.0 (72.0, 95.7) | 80.0 (70.0, 90.0) | 79.2 (67.6, 91.1) | < 0.001 |
| BMI, median (IQR), kg/m^2^ | 28.3 (25.0, 32.2) | 26.3 (24.2, 29.4) | 27.4 (24.3, 30.6) | < 0.001 |
| Gender (Number & Percentage of Males) | 4773 (71.85%) | 1642 (72.85%) | 597 (70.57%) | < 0.001 |
| Maximum SOFA Score, median (IQR) | 6.0 (4.0, 8.0) | 6.0 (6.0, 8.0) | 10.0 (9.0, 12.0) | < 0.001 |
| Base Excess, median (IQR), mEq/L | -1.0 (-2.0, 0.0) | -0.6 (-2.5, 1.0) | -1.1 (-2.9, 0.7) | < 0.001 |
| PO_2_, median (IQR), mmHg | 117.0 (94.0, 155.0) | 86.4 (75.4, 101.8) | 95.0 (80.0, 118.0) | < 0.001 |
| SO_2_, median (IQR), percentage | 97.0 (95.0, 98.0) | 96.4 (95.0, 97.5) | 98.0 (97.0, 99.0) | < 0.001 |
| PCO_2_, median (IQR), mmHg | 39.5 (36.0, 43.0) | 38.1 (35.1, 41.5) | 40.0 (36.0, 43.0) | < 0.001 |
| Chloride, median (IQR), mEq/L | 107.0 (105.0, 109.0) | 105.0 (103.0, 107.0) | 107.0 (104.0, 109.0) | < 0.001 |
| Total CO_2_, median (IQR), mEq/L | 24.0 (23.0, 26.0) | 22.6 (22.1, 23.4) | 25.0 (23.0, 27.0) | < 0.001 |
| pH, median (IQR) | 7.38 (7.35, 7.41) | 7.41 (7.38, 7.44) | 7.38 (7.35, 7.42) | < 0.001 |
| PaO_2_ Fio_2_ Ratio, median (IQR), mmHg | 257.5 (196.0, 330.0) | 251.0 (194.2, 324.6) | 232.5 (161.7, 316.1) | < 0.001 |
| FiO_2_, median (IQR) | 50.0 (40.0, 50.0) | 40.0 (35.0, 47.0) | 40.0 (40.0, 60.0) | < 0.001 |
| Alveolar-arterial oxygen gradient, median (IQR), mmHg | 170.2 (122.0, 222.0) | 134.6 (91.9, 186.9) | 147.4 (110.2, 244.8) | < 0.001 |
| Lactate, median (IQR), mmol/L | 1.9 (1.4, 2.5) | 1.3 (1.0, 1.7) | 1.2 (0.9, 1.7) | < 0.001 |
| Sodium, median (IQR), mEq/L | 137.0 (135.0, 139.0) | 137.0 (134.3, 139.0) | 136.0 (134.0, 139.0) | < 0.001 |
| Potassium, median (IQR), mEq/L | 4.3 (4.0, 4.6) | 4.3 (4.1, 4.6) | 4.4 (4.1, 4.6) | < 0.001 |
| Bicarbonate, median (IQR), mEq/L | 24.0 (22.0, 26.0) | 23.7 (22.0, 25.4) | 23.2 (21.0, 24.8) | < 0.001 |
| Calcium, median (IQR), mg/dL | 8.8 (8.3, 9.1) | 8.1 (7.8, 8.4) | 7.8 (7.6, 8.1) | < 0.001 |
| Glucose, median (IQR), mg/dL | 124.3 (108.0, 144.0) | 130.0 (120.8, 151.4) | 146.0 (124.0, 172.0) | < 0.001 |
| BUN, median (IQR), mg/dL | 17.0 (13.0, 22.7) | 14.5 (11.7, 19.6) | 16.0 (12.0, 21.0) | < 0.001 |
| Anion gap, median (IQR), mEq/L | 12.0 (10.0, 14.0) | 10.4 (8.5, 12.4) | 8.3 (6.8, 10.3) | < 0.001 |
| Creatinine, median (IQR), mg/dL | 0.90 (0.80, 1.20) | 0.89 (0.72, 1.10) | 0.86 (0.70, 1.06) | < 0.001 |
| Baseline Creatinine, median (IQR), mg/dL | 0.90 (0.80, 1.20) | 0.92 (0.80, 1.10) | 0.88 (0.74, 1.09) | < 0.001 |
| Hematocrit, median (IQR), percentage | 29.6 (27.0, 32.5) | 28.0 (25.0, 31.5) | 29.0 (26.1, 33.0) | < 0.001 |
| Hemoglobin, median (IQR), g/dL | 10.0 (9.1, 11.0) | 9.6 (8.5, 10.7) | 9.7 (8.7, 11.0) | < 0.001 |
| WBC, median (IQR), ×10³/µL | 12.5 (9.9, 15.9) | 11.0 (9.0, 13.8) | 12.5 (10.1, 15.6) | < 0.001 |
| RBC, median (IQR), ×10⁶/µL | 3.3 (3.0, 3.7) | 3.1 (2.8, 3.5) | 3.3 (2.9, 3.7) | < 0.001 |
| RDW, median (IQR), percentage | 14.0 (13.2, 15.0) | 14.2 (13.7, 14.5) | 13.6 (13.0, 14.7) | < 0.001 |
| MCH, median (IQR), pg | 30.4 (29.3, 31.5) | 30.5 (29.5, 31.6) | 30.0 (28.7, 31.3) | < 0.001 |
| MCHC, median (IQR), g/dL | 33.7 (32.8, 34.6) | 34.2 (33.5, 34.9) | 33.2 (32.5, 34.1) | < 0.001 |
| MCV, median (IQR), fL | 90.0 (87.0, 93.0) | 89.1 (86.3, 92.0) | 89.9 (86.4, 93.4) | < 0.001 |
| Platelet, median (IQR), ×10³/µL | 143.0 (113.0, 180.0) | 146.0 (116.0, 174.0) | 140.5 (113.0, 169.0) | < 0.001 |
| PT, median (IQR), seconds | 13.6 (12.7, 14.7) | 9.1 (8.0, 10.7) | 13.8 (13.2, 15.3) | < 0.001 |
| PTT, median (IQR), seconds | 30.1 (27.4, 34.4) | 36.0 (33.0, 40.0) | 33.2 (30.2, 37.6) | < 0.001 |
| INR, median (IQR) | 1.2 (1.2, 1.3) | 1.2 (1.2, 1.2) | 1.3 (1.2, 1.4) | < 0.001 |
| Diastolic Blood Pressure, median (IQR), mmHg | 57.0 (51.0, 62.8) | 55.0 (49.9, 61.1) | 56.0 (51.0, 62.0) | < 0.001 |
| Mean Arterial Pressure, median (IQR), mmHg | 73.0 (67.0, 80.0) | 72.7 (66.8, 80.4) | 76.0 (70.0, 83.0) | < 0.001 |
| Systolic Blood Pressure, median (IQR), mmHg | 111.0 (102.0, 122.0) | 112.0 (101.0, 125.4) | 118.0 (107.0, 129.5) | < 0.001 |
| Heart Rate, median (IQR), beats/min | 82.0 (75.0, 90.0) | 80.0 (74.6, 87.4) | 80.5 (73.0, 90.5) | < 0.001 |
| Respiratory rate, median (IQR), breaths/min | 18.8 (15.8, 22.0) | 15.0 (14.0, 16.4) | 18.2 (15.0, 22.0) | < 0.001 |
| Temperature, median (IQR), 75% IQR), Celsius | 37.0 (36.7, 37.2) | 37.3 (37.0, 37.6) | 36.8 (36.4, 37.3) | < 0.001 |
| SPO_2_, median (IQR), percentage | 97.0 (95.0, 99.0) | 97.1 (95.4, 98.7) | 97.0 (95.0, 99.0) | < 0.001 |
| ALP, median (IQR), U/L | 81.0 (71.0, 91.9) | 72.0 (62.0, 89.9) | 46.0 (39.0, 57.0) | < 0.001 |
| AST, median (IQR), U/L | 44.7 (31.2, 66.0) | 38.9 (31.1, 90.8) | 61.0 (42.0, 90.0) | < 0.001 |
| ALT, median (IQR), U/L | 40.5 (27.0, 68.9) | 42.0 (28.3, 92.4) | 21.0 (15.0, 31.0) | < 0.001 |
| Bilirubin Total, median (IQR), mg/dL | 0.6 (0.6, 0.8) | 0.6 (0.6, 0.7) | 0.8 (0.6, 1.2) | < 0.001 |
| Fluid Balance, median (IQR), ml | -20.3 (-74.9, 51.2) | 37.1 (-15.7, 127.3) | -4.9 (-55.0, 50.0) | < 0.001 |
| Cumulative Fluid Balance, median (IQR), ml | 3630.1 (1976.0, 5228.4) | 3262.0 (1602.6, 5480.8) | 1991.2 (736.5, 3241.1) | < 0.001 |
| Urine Output Amount, median (IQR), ml | 62.0 (35.5, 122.2) | 50.0 (31.6, 79.2) | 50.0 (30.0, 100.0) | < 0.001 |
| Nephrotoxins - (Number & Percentage of Doses Given) | 2724 (41.01%) | 308 (13.66%) | 821 (97.04%) | < 0.001 |

**Abbreviations**: MIMIC IV, Medical Information Mart for Intensive Care (MIMIC)-IV; SICdb, Salzburg Intensive Care database; MSHS: Mount Sinai Hospital System; IQR: Inter Quartile Range; cm, Centimeters; kg, Kilograms; BMI, Body Mass Index; kg/m², Kilograms per Square Meter; SOFA, Sequential Organ Failure Assessment; PO₂, Partial pressure of Oxygen; mEq/L, Milliequivalents per Liter; mmHg, Millimeters of Mercury; SO₂, Oxygen Saturation; PCO₂, Partial Pressure of Carbon Dioxide; CO₂, Carbon Dioxide; PH, Potential of Hydrogen; PaO₂/FiO₂, Arterial Oxygen Partial Pressure to Fraction of Inspired Oxygen Ratio; mg/dL, Milligrams per Deciliter; FiO₂, Fraction of Inspired Oxygen; BUN, Blood Urea Nitrogen; RBC, Red Blood Cell; WBC, White Blood Cell, RDW, Red Cell Distribution Width; MCH, Mean corpuscular hemoglobin; MCHC, Mean Corpuscular Hemoglobin Concentration; MCV, Mean Corpuscular Volume; PT, Prothrombin Time; PTT, Partial Thromboplastin Time; INR, International Normalized Ratio; SpO₂, Peripheral Capillary Oxygen Saturation; ALP, Alkaline Phosphatase; U/L, Units per Liter; AST, Aspartate Aminotransferase; ALT, Alanine Aminotransferase; ml, Milliliters.

**e**-**Figure 1****:** CONSORT Diagram for inclusion and exclusion criteria of the study

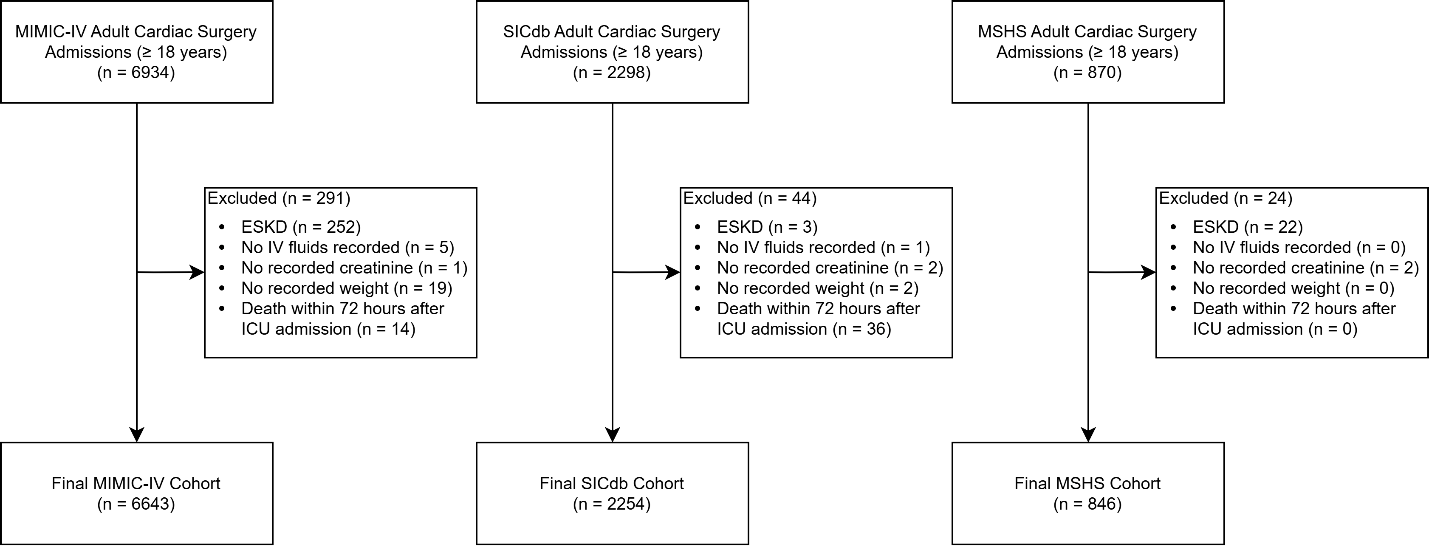

**Abbreviations**: MIMIC IV, Medical Information Mart for Intensive Care (MIMIC)-IV; SICdb, Salzburg Intensive Care database; MSHS: Mount Sinai Hospital System; ESKD, End-Stage Kidney Disease; IV Fluids, Intravenous Fluids; ICU, Intensive Care Unit.

**e**-**Figure 2-a:** Reinforcement Learning vs. Clinician Actions Comparison (Time)

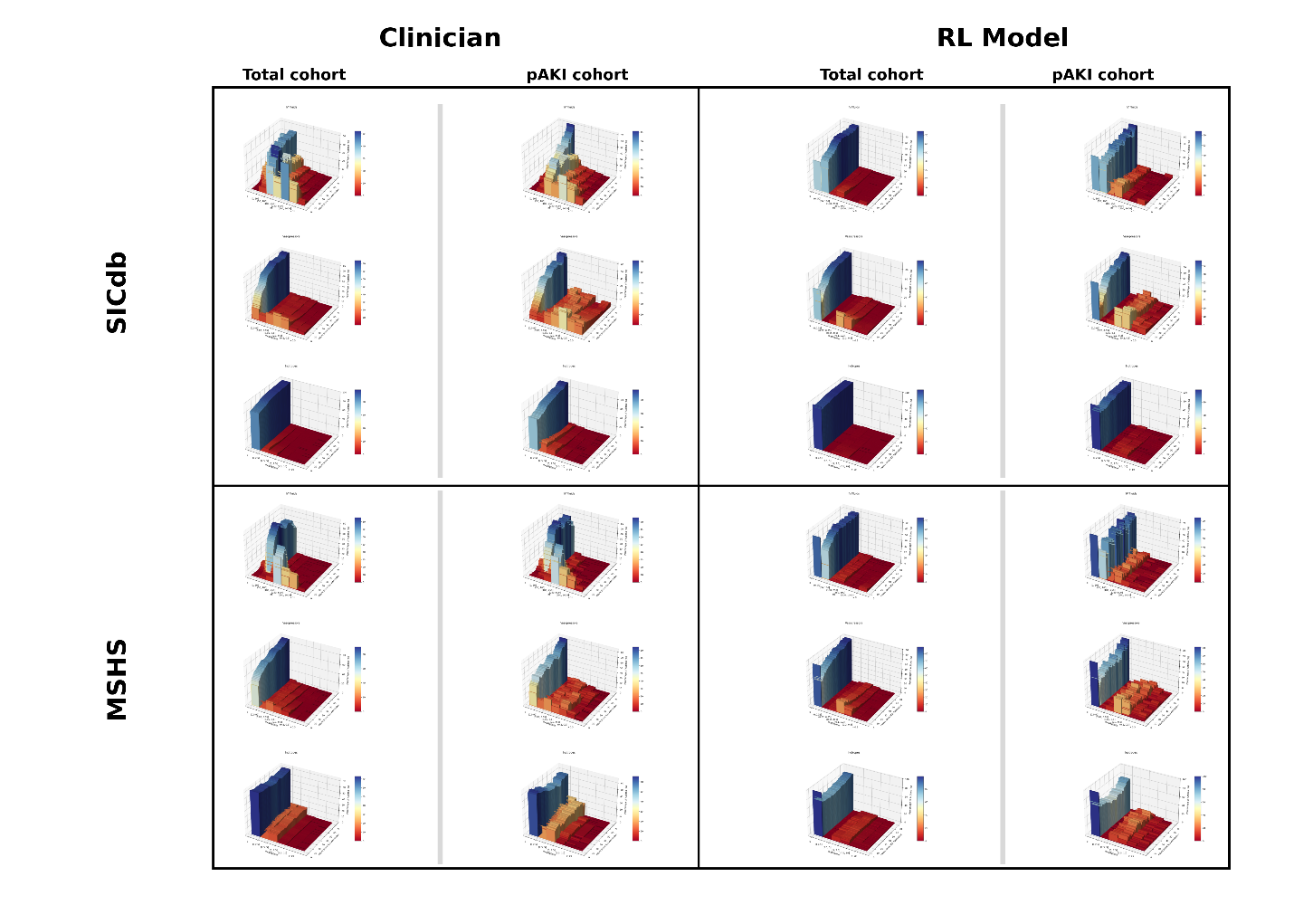

**e**-**Figure 2-b:** Reinforcement Learning vs. Clinician Actions Comparison (MAP)

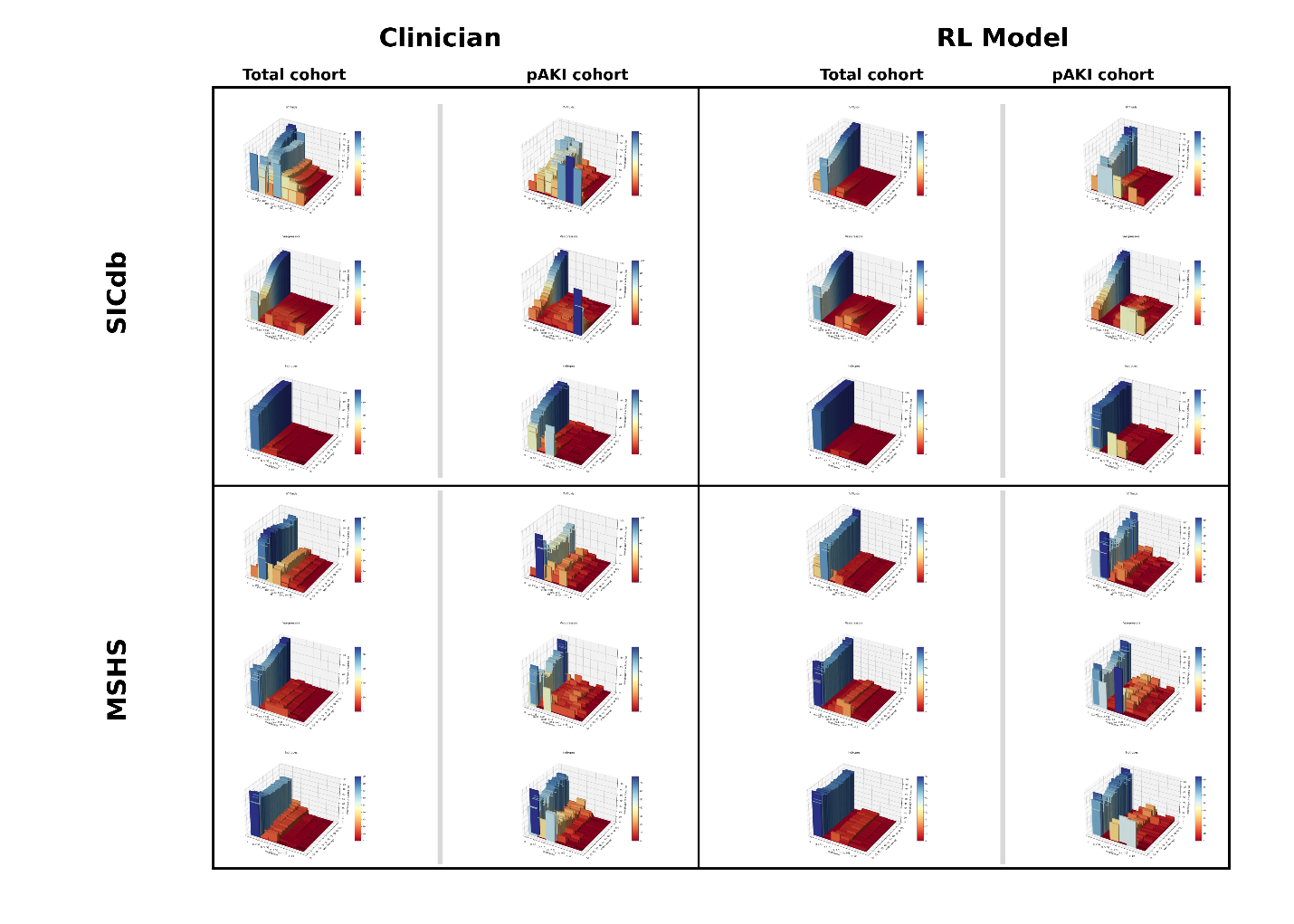

**Abbreviations**: SICdb, Salzburg Intensive Care database; MSHS: Mount Sinai Hospital System; IV Fluids, Intravenous Fluids; ICU, Intensive Care Unit; pAKI, persistent Acute Kidney Injury; MAP, Mean Arterial Pressure; mmHg, Millimeters of Mercury; ml, Milliliters; mcg/kg/min, micrograms per kilogram per minute.

**e**-**Figure** **3-a: SICdb** Patient No. 6 Hourly Top Ten SHAP Values Heatmap

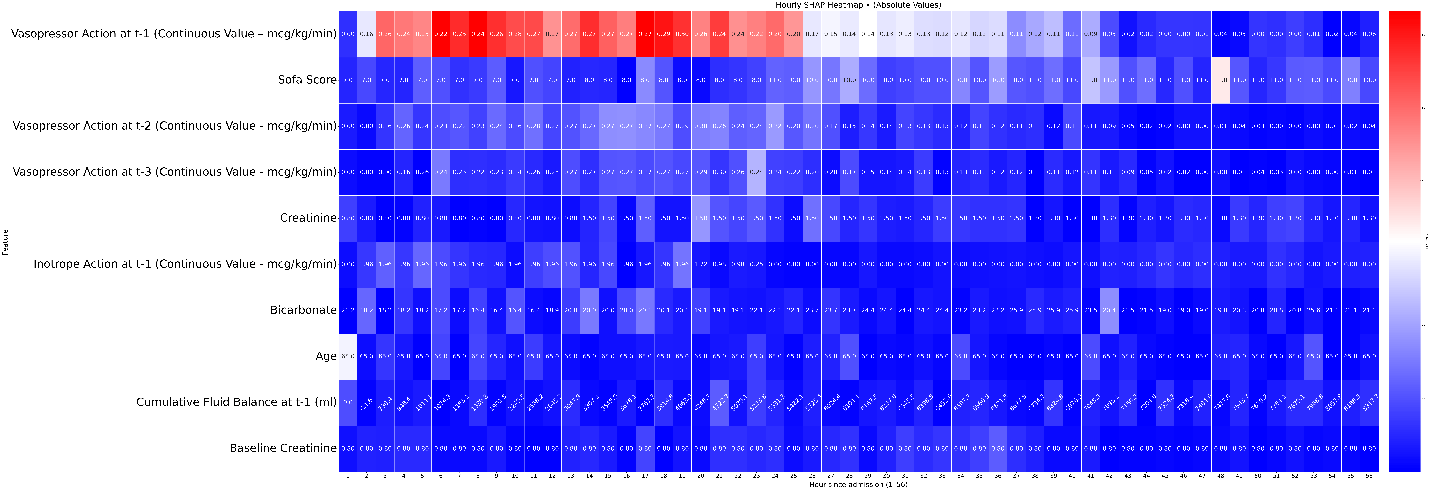

**e**-**Figure** **3-b: SICdb** Patient No. 10 Hourly Top Ten SHAP Values Heatmap

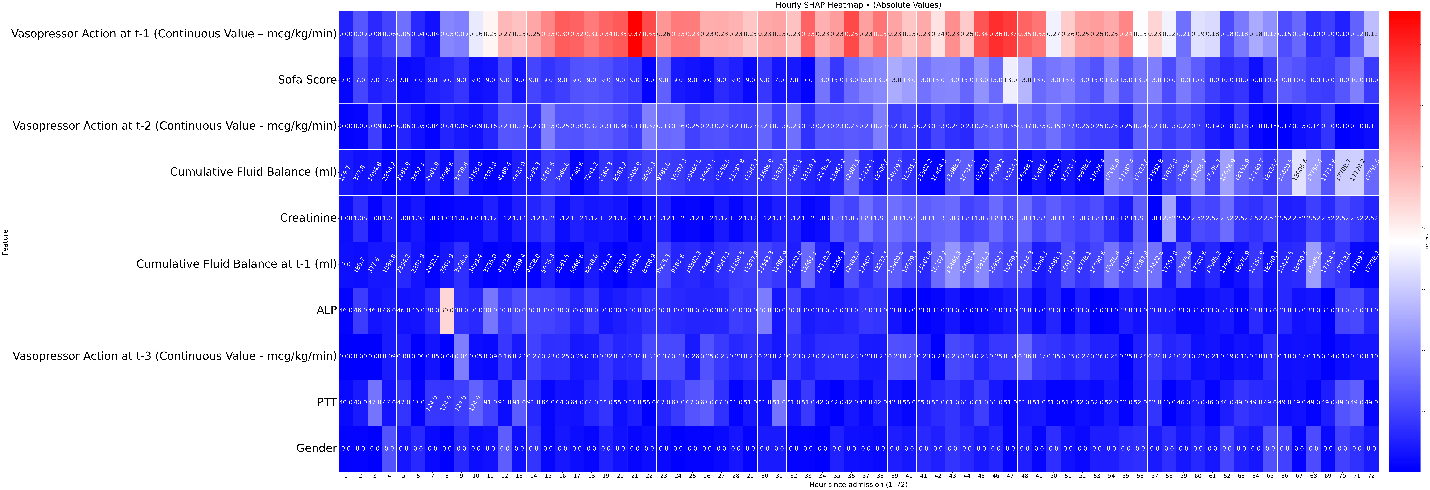

**e**-**Figure** **3-c: SICdb** Patient No. 12 Hourly Top Ten SHAP Values Heatmap

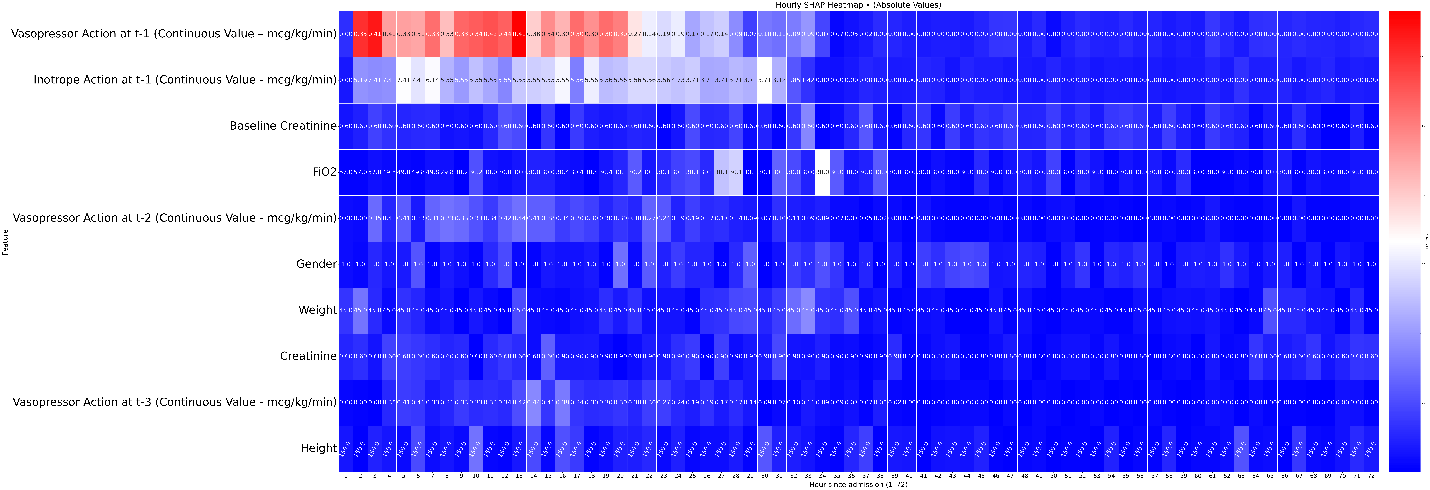

**e**-**Figure** **3-d: SICdb** Patient No. 19 Hourly Top Ten SHAP Values Heatmap

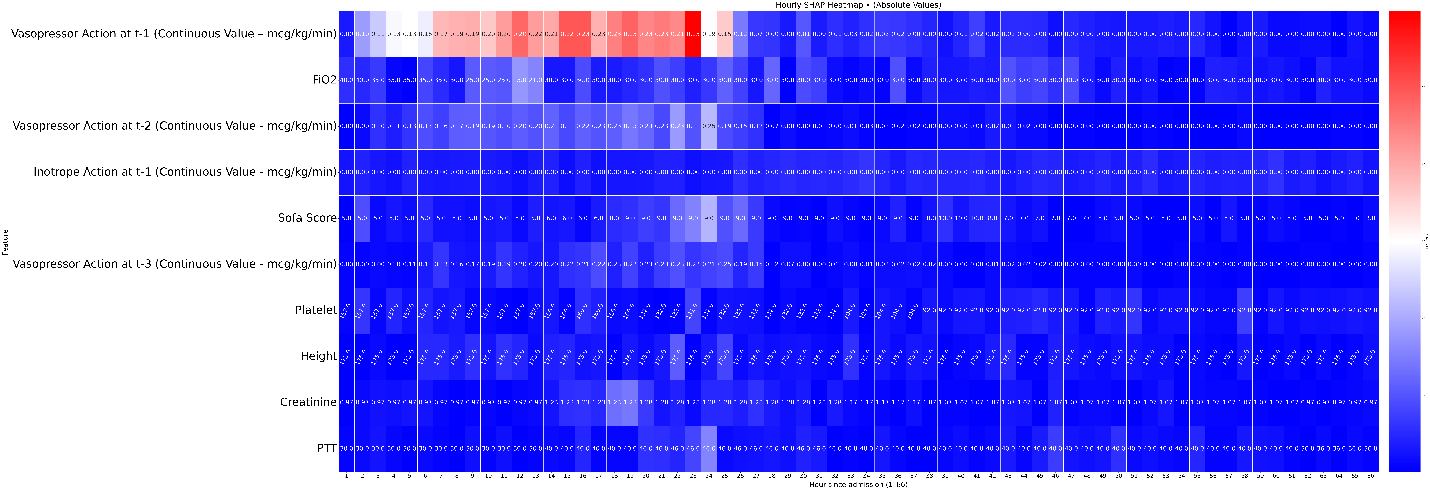

**e**-**Figure** **3-e: SICdb** Patient No. 30 Hourly Top Ten SHAP Values Heatmap

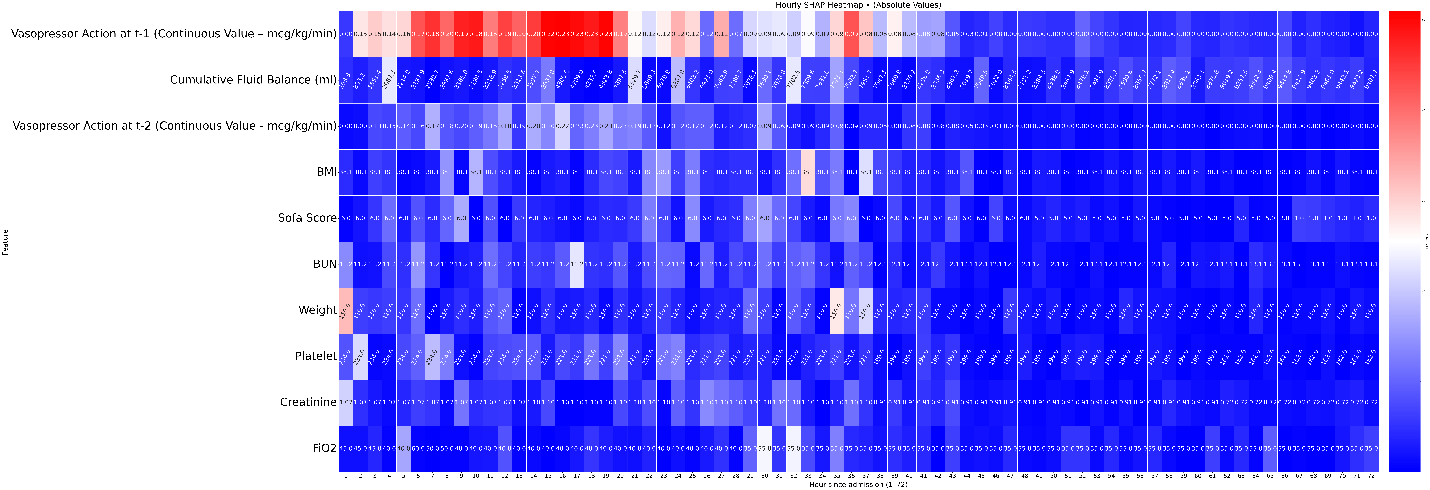

**e**-**Figure** **3-f: MSHS** Patient No. 6 Hourly Top Ten SHAP Values Heatmap

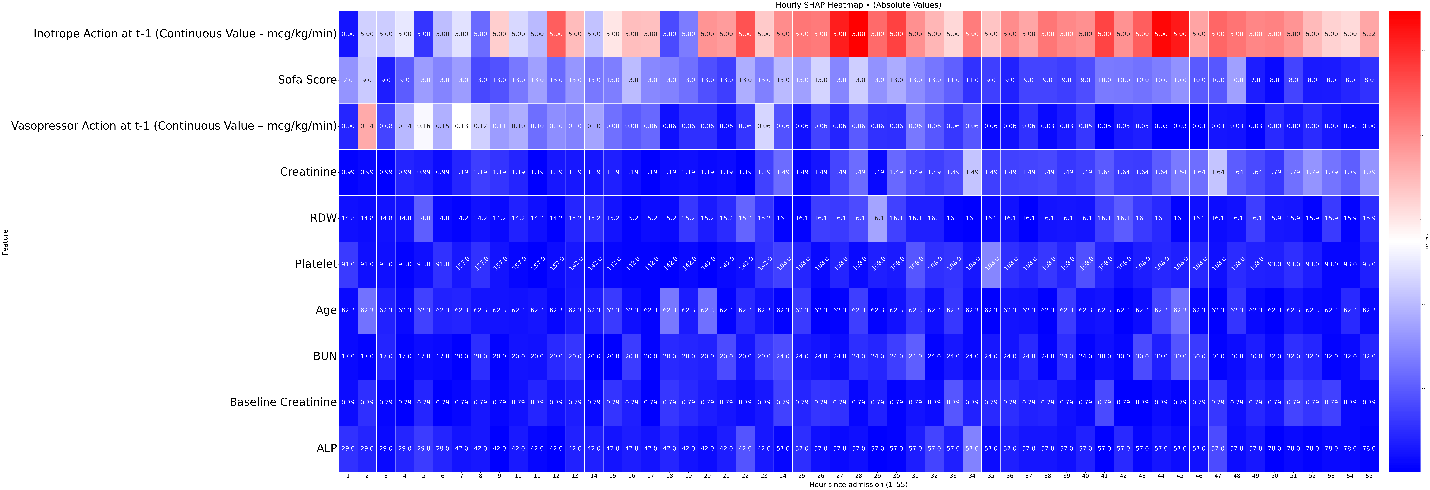

**e**-**Figure** **3-g: MSHS** Patient No. 10 Hourly Top Ten SHAP Values Heatmap

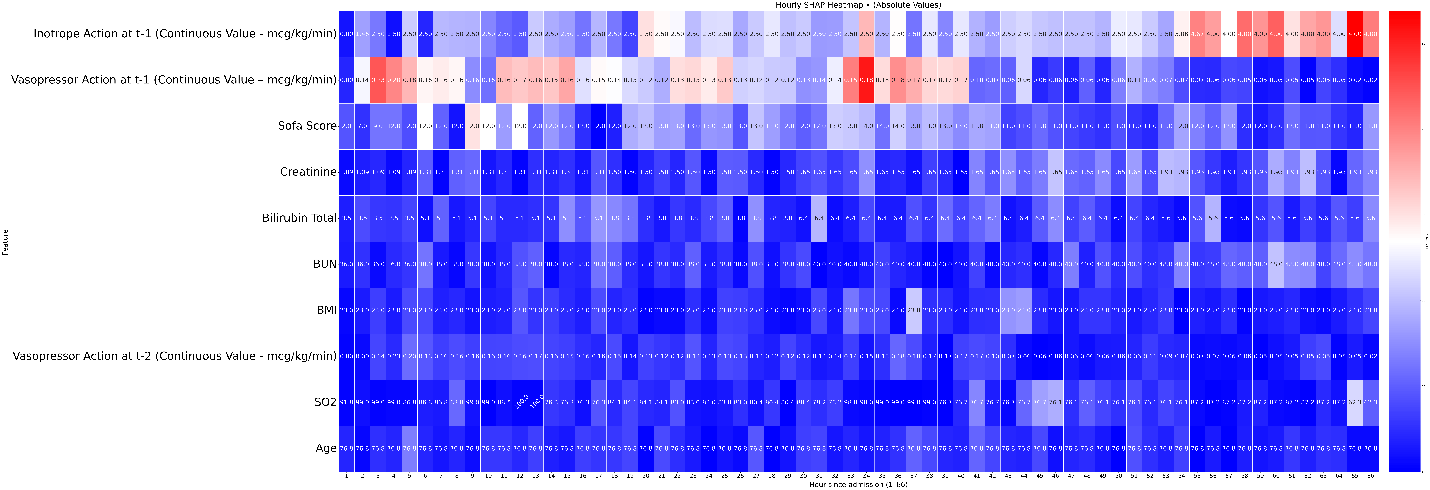

**e**-**Figure** **3-h: MSHS** Patient No. 12 Hourly Top Ten SHAP Values Heatmap

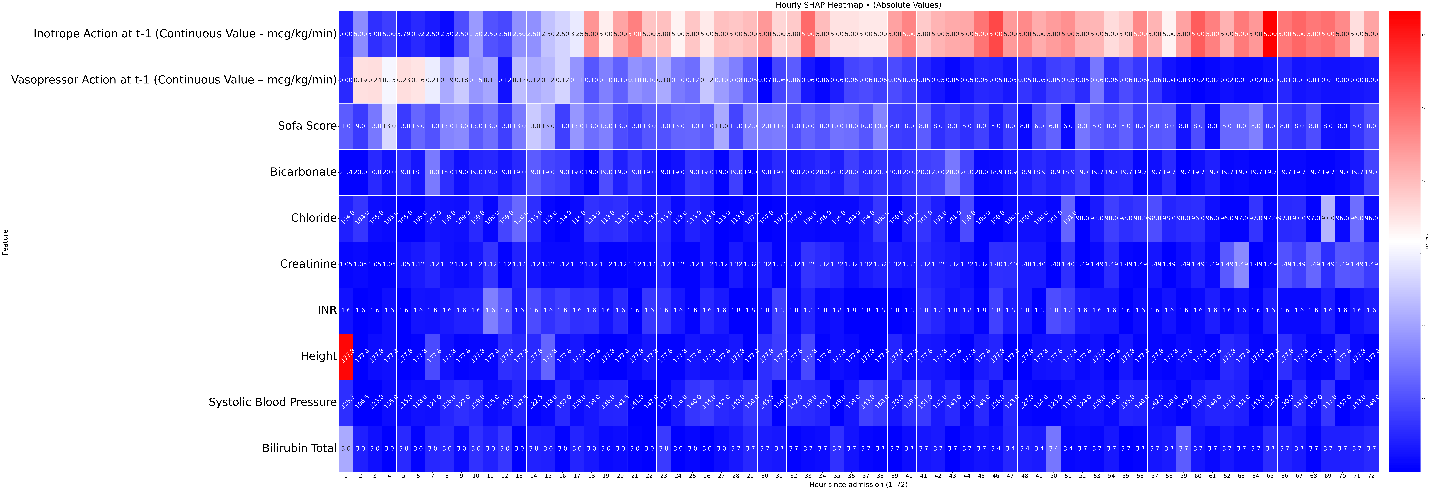

**e**-**Figure** **3-i: MSHS** Patient No. 19 Hourly Top Ten SHAP Values Heatmap

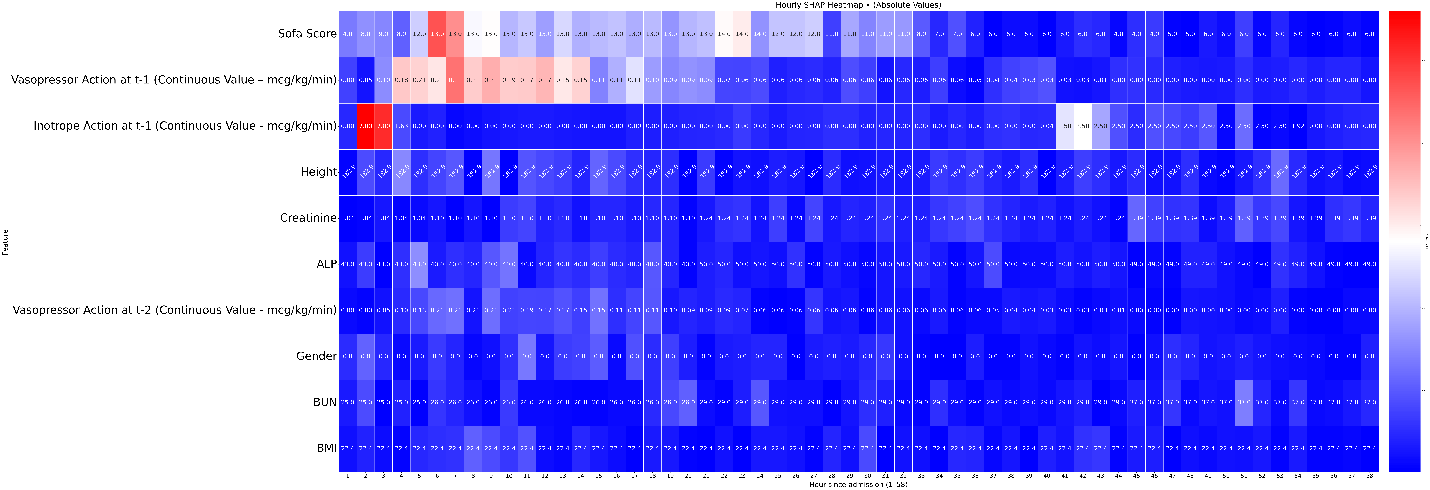

**e**-**Figure** **3-j: MSHS** Patient No. 30 Hourly Top Ten SHAP Values Heatmap

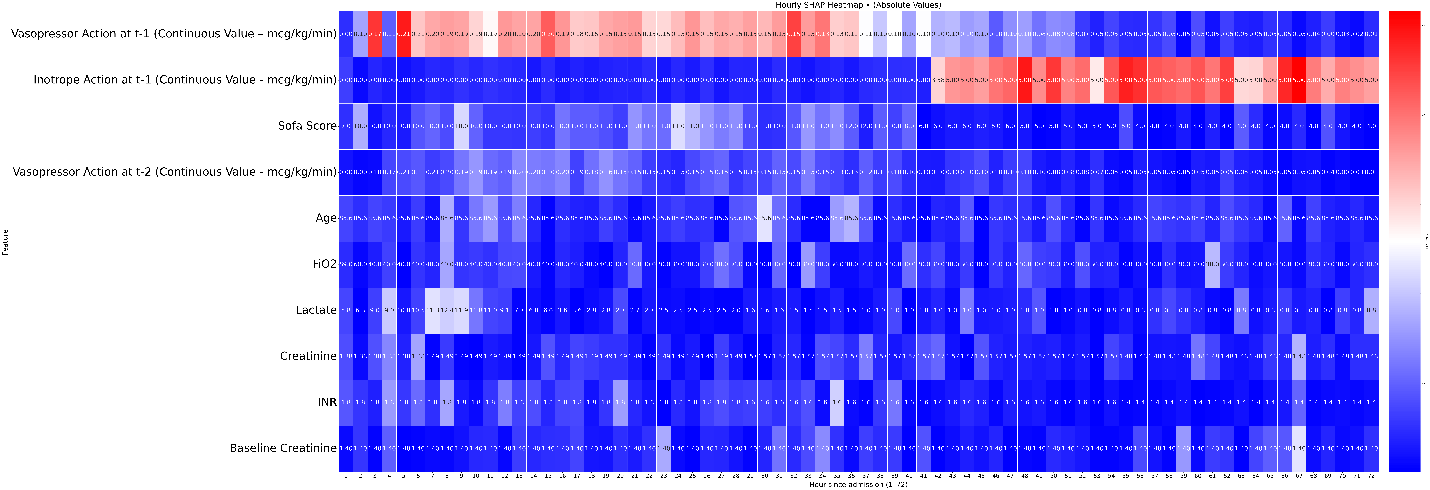

**Abbreviations**: SICdb, Salzburg Intensive Care database; MSHS: Mount Sinai Hospital System; IV Fluids, Intravenous Fluids; ICU, Intensive Care Unit; pAKI, persistent Acute Kidney Injury; MAP, Mean Arterial Pressure; ml, Milliliters; mcg/kg/min, micrograms per kilogram per minute; SOFA, Sequential Organ Failure Assessment; ALP, Alkaline Phosphatase; PTT, Partial Thromboplastin Time; FiO₂, Fraction of Inspired Oxygen; BMI, Body Mass Index; BUN, Blood Urea Nitrogen; RDW, Red Cell Distribution Width; SO₂, Oxygen Saturation; INR, International Normalized Ratio; t-1, previous hour value; t-2, Value in two hours ago; t-3: Value in three hours ago.
